# Cross-sectional study of the association between depressive symptoms and attentional bias to emotional stimuli in patients with acute stroke: Study protocol

**DOI:** 10.64898/2026.06.13.26355568

**Authors:** Miyu Yamashita, Hirokazu Takizawa, Kohei Koizumi, Toyohiro Hamaguchi

## Abstract

Post-stroke depression affects approximately 30% of patients after stroke and is associated with delayed recovery in activities of daily living, reduced rehabilitation effectiveness, and poorer quality of life. Attentional bias modification may provide a low-burden, nonpharmacological approach for patients in the acute phase of stroke. However, before such an intervention can be implemented in clinical practice, it is necessary to clarify whether attentional bias is present in patients with acute stroke and depressive symptoms, whether cognitive function influences the manifestation of this bias, and which task and stimulus formats are most appropriate for assessment. This multicenter, cross-sectional observational study will enroll patients with acute stroke between 7–30 days after stroke onset. Depressive symptoms will be assessed using the depression subscale of the Hospital Anxiety and Depression Scale. Attentional bias will be measured under four task conditions based on the dot-probe task and the cue–target task, using face and word stimuli. Secondary assessments will include cognitive function, anxiety symptoms, activities of daily living, health-related quality of life, and clinical background variables. The aims of this study are to investigate the association between depressive symptoms and attentional bias in patients with acute stroke, compare attentional bias characteristics across task and stimulus types, and examine the potential influence of cognitive function on this association. The findings are expected to provide an empirical basis for designing future attentional bias modification protocols targeting post-stroke depression in the acute phase. This study has been registered with the UMIN Clinical Trials Registry (UMIN000059166).

## Introduction

Stroke remains a major global health challenge and is one of the leading causes of disability worldwide [1]. In addition to motor, sensory, and cognitive impairments, many patients experience psychological complications following stroke. Among these, post-stroke depression (PSD) is one of the most prevalent and clinically important conditions, affecting approximately 30% of stroke survivors [2]. PSD has been associated with delayed recovery in activities of daily living (ADL), poorer functional independence, and reduced quality of life [3,4]. The prevalence of PSD is particularly high, with approximately 30% reported within one month of stroke onset [5], and early PSD has been associated with persistent depressive symptoms and functional impairment [6]. These findings suggest that the acute phase after stroke may represent a critical window for early identification and intervention.

Despite its clinical significance, effective and feasible early interventions for depressive symptoms in patients with acute stroke have not been firmly established. Pharmacological treatment options may be limited by concerns regarding adverse events, including seizures, falls, and delirium [7]. Nonpharmacological approaches, such as cognitive behavioral therapy, may be beneficial [8]; however, participation in structured psychotherapy may be difficult during the acute phase because patients often experience physical symptoms, fatigue, reduced concentration, and cognitive impairment. Therefore, there is a clinical need for a noninvasive, low-burden, and safe psychological approach that can be implemented early after stroke.

Attention bias modification (ABM) may offer one such approach. ABM is based on the premise that depression is associated with maladaptive attentional processing of emotional information, including selective attention to negative stimuli and difficulty disengaging attention from such stimuli [9,10]. Because ABM can be delivered using brief computerized tasks and does not require extensive verbal processing or introspection, it may be particularly suitable for patients in the acute stage after stroke. In non-stroke populations, randomized controlled trials and meta-analyses have suggested that ABM may alleviate depressive symptoms [11,12].

However, its applicability to patients with acute stroke remains unclear.

Before ABM can be appropriately adapted for acute stroke populations, several questions must be addressed. First, it remains unknown whether attentional bias is present in the acute phase after stroke and how strongly it is associated with depressive symptoms. In patients in the convalescent stage after stroke, Takizawa et al. found that attentional orienting was related to cognitive function rather than depressive symptoms. Specifically, among patients with depressive symptoms, those without mild cognitive impairment (MCI) selected neutral facial expressions more rapidly than those with MCI, indicating preferential orienting toward neutral rather than aversive faces; depressive symptoms alone did not significantly affect performance [13]. However, the acute phase differs substantially from the convalescent phase because early cognitive impairment is common after stroke [14], and difficulties in emotion regulation may also influence attentional processing [15]. In healthy older adults, the positivity effect—preferential attention to positive rather than negative information—has been linked to cognitive control [16,17]. Accordingly, reduced cognitive resources in acute stroke may alter patterns of attentional bias, and the characteristics of attentional bias in this phase should be examined directly rather than inferred from later-stage populations.

Second, it remains unclear which task format is most appropriate for evaluating attentional bias in patients with acute stroke. Two candidate paradigms are the dot-probe task (DPT) and the cue–target task (CTT). These paradigms may capture different components of attentional processing. Specifically, the DPT has been used primarily to assess selective orienting toward threatening or negative stimuli [18], whereas the CTT may more effectively reflect difficulty disengaging attention from emotionally salient stimuli [19]. In subthreshold depression, difficulty disengaging attention has been reported even when clear evidence of selective orienting was not observed [20]. Therefore, task-related differences in attentional bias may be especially relevant when considering the optimal design of ABM interventions for this population.

Third, the optimal stimulus format for assessing attentional bias in acute stroke is unclear. Emotional faces and emotional words are likely to engage partially distinct processing pathways. Facial expressions may be more strongly related to externally driven social-emotional attention [21], whereas emotional words may reflect more internally mediated semantic and self-referential processing [22]. In older adults with and without major depressive disorder, Ros et al. suggested that emotional words and emotional faces may be processed differently [23]. It is therefore plausible that patients with acute stroke may also exhibit different patterns of attentional bias depending on stimulus modality. Clarifying these differences is important for selecting the most appropriate stimuli for future ABM paradigms.

Taken together, there is a clear knowledge gap regarding attentional bias in patients with acute stroke. Although PSD is common in the early phase after stroke and ABM may represent a promising low-burden intervention strategy, the fundamental characteristics of attentional bias in this population have not yet been sufficiently characterized. In particular, uncertainty remains regarding the association between depressive symptoms and attentional bias, the relative utility of different task formats, the influence of stimulus type, and the potential moderating role of cognitive function. Addressing these issues is necessary before developing and testing ABM interventions tailored to patients with acute stroke.

The primary objective of this multicenter cross-sectional observational study is to investigate the association between depressive symptoms and attentional bias indices in patients with acute stroke. The secondary objectives are to compare attentional bias patterns across task formats (DPT vs CTT) and stimulus types (facial vs word stimuli) and to explore whether cognitive function moderates the relationship between depressive symptoms and attentional bias. These findings are expected to provide an empirical foundation for designing future attentional bias modification protocols, including participant selection criteria, task format, and stimulus format. The schedule of participant enrollment and assessments is presented in Fig 1. An overview of the study rationale and design is presented in Fig 2.

**Fig 1.**
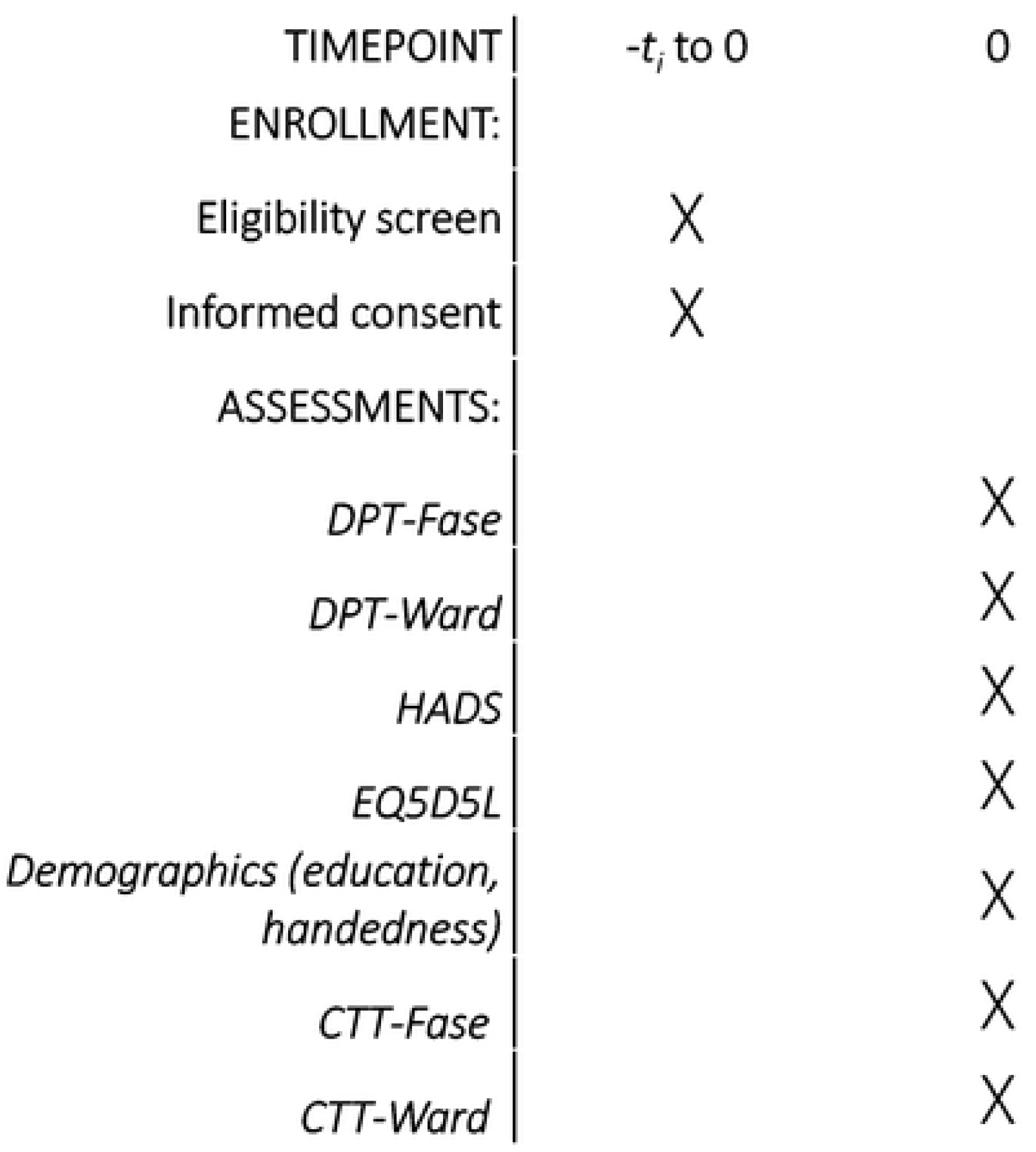
Schedule of enrollment and assessments. Schedule of participant enrollment, eligibility screening, informed consent, and assessment procedures, including demographic information, Hospital Anxiety and Depression Scale–Depression, EuroQol 5 Dimensions 5 Levels, and attentional bias measures using the DPT and CTT.

**Fig 2.**
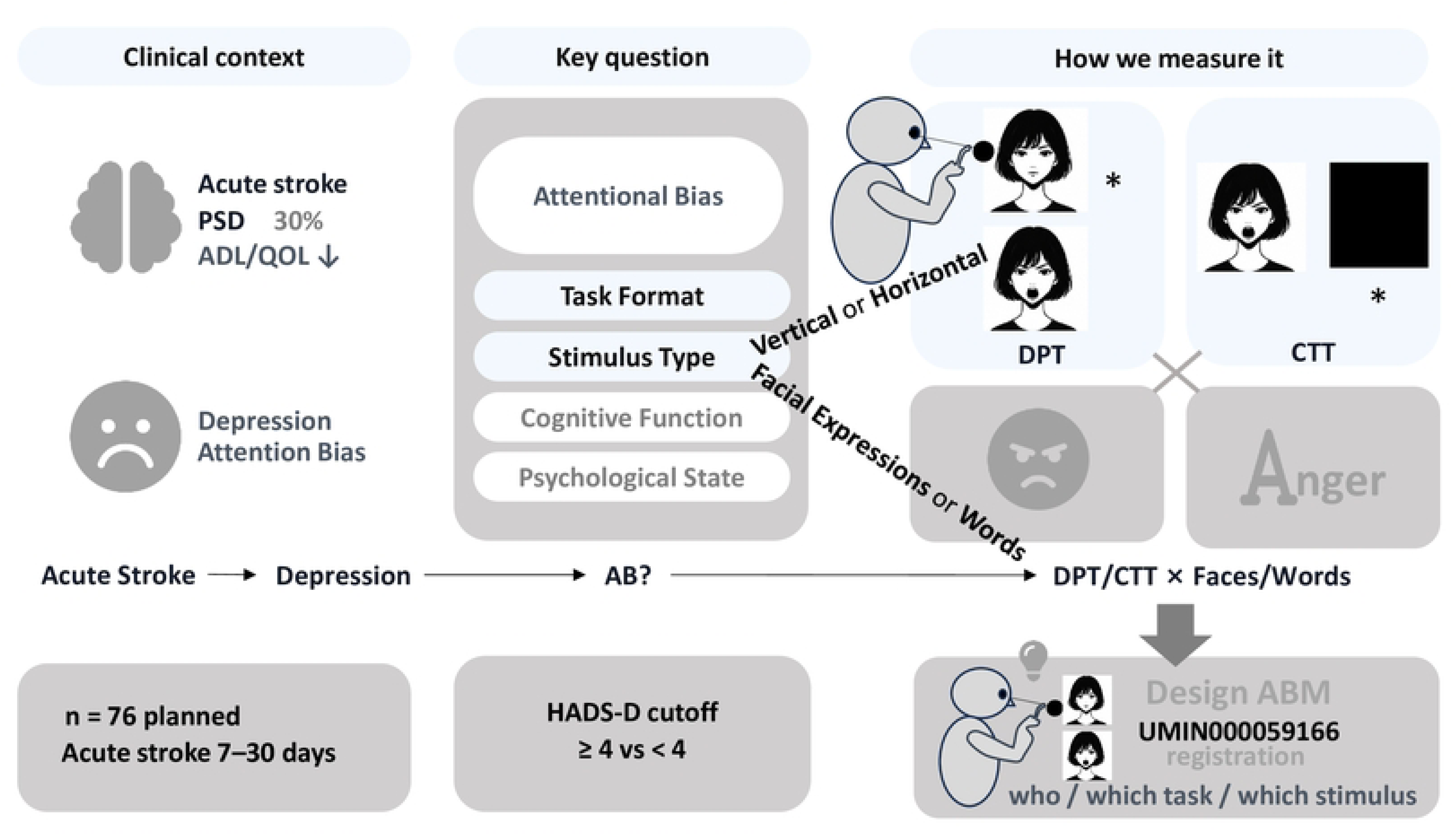
Graphical abstract. Post-stroke depression (PSD) affects approximately 30% of stroke survivors and is associated with adverse effects on activities of daily living and quality of life. A substantial proportion of depressive symptoms emerges within the first month after stroke onset. Attentional bias, including selective attention to negative information and difficulty disengaging attention from negative stimuli, has been implicated in depression. The present protocol describes a multicenter cross-sectional observational study that will recruit patients 7–30 days after stroke onset and will assess depressive symptoms, anxiety symptoms, quality of life, and attentional bias using the dot-probe task and the cue–target task with facial and word stimuli. The planned sample size is 76 participants. The findings are expected to inform the design of future ABM intervention protocols, including target participant selection, task format, and stimulus type. The image stimuli used in this paper are illustrative and do not represent the actual images used in the study.

## Materials and methods

### Study design and setting

This study is a multicenter cross-sectional observational study designed to examine the association between depressive symptoms and attentional bias to emotional stimuli in patients with acute stroke. The study will be conducted as a collaborative project involving Niiza Shiki Central General Hospital, Nishitokyo Chuo General Hospital, Totsuka Kyoritsu Izumino Hospital, and Saitama Prefectural University. Participant recruitment and study assessments will be conducted at the participating hospitals, and data analysis will be performed by the research team in collaboration with the university. The participant screening, exclusion, and group classification procedures are summarized in Fig 2.

### Participants

The study population will consist of hospitalized patients with acute stroke, including cerebral infarction and intracerebral hemorrhage, who are receiving inpatient care at the participating hospitals. Patients will be eligible if they meet all of the following criteria: physician-diagnosed stroke; prescription for occupational therapy; 7–30 days after stroke onset; age 20 years or older; a Mini-Mental State Examination (MMSE) score of 24 or higher; and a level of consciousness within the one-digit range on the Japan Coma Scale. Patients will be excluded if they have subarachnoid hemorrhage, severe visual impairment, a history of depression prior to stroke onset, a first-degree family history of severe depression, a serious comorbid illness, or severe aphasia that substantially interferes with communication and task performance.

### Recruitment and informed consent

Due to clinical and operational constraints, consecutive screening of all admitted patients will not be performed. Instead, screening will be conducted among patients admitted to the participating hospitals during the study period who are accessible for assessment and follow-up by the co-investigators. This selection method may introduce potential selection bias. For patients judged to meet the eligibility criteria, a co-investigator will provide a detailed explanation of the study using a written information sheet. After confirming the participant’s full understanding, written informed consent will be obtained. The numbers of patients screened, excluded (due to ineligibility or refusal), and excluded from analysis will be recorded. Once eligibility is confirmed, the co-investigators will proceed with the explanation and obtain consent.

### Study procedures

After written informed consent is obtained, participants will undergo a study assessment lasting approximately 40 minutes during routine occupational therapy hours. The assessment may be completed in a single session, divided into multiple sessions, or postponed or discontinued, depending on the participant’s physical condition and fatigue level. Assessments will be conducted in a private room whenever possible; if this is not feasible due to clinical risk management considerations, the assessment will be conducted at the bedside with the curtain closed. The assessment environment will be recorded.

The planned assessment sequence is as follows: preparation, including a vital sign check, postural adjustment, and task explanation (approximately 5 minutes); practice and assessment for the dot-probe task with facial stimuli (DPT-Face) and the dot-probe task with word stimuli (DPT-Word) (approximately 4 minutes); psychological assessments, including depressive symptoms, anxiety symptoms, and health-related quality of life (approximately 15 minutes); collection of handedness and educational history, with rest provided as needed (approximately 5 minutes); practice and assessment for the cue–target task with facial stimuli (CTT-Face) and the cue–target task with word stimuli (CTT-Word) (approximately 6 minutes); and a post-assessment vital sign check (approximately 5 minutes). The total assessment time will be approximately 40 minutes. If a participant’s physical condition does not permit safe assessment on the scheduled day, the assessment may be postponed.

To ensure adequate task performance, each attentional bias task will include a practice mode consisting of 16 trials. If the accuracy rate in the first 10 practice trials is below 70%, the participant will be allowed to repeat the practice session up to three times. Participants who do not reach the predefined accuracy threshold will remain enrolled in the study but will be excluded from the corresponding attentional bias analyses [24].

Standard rehabilitation, including physical therapy, occupational therapy, and speech-language therapy, will be permitted during the study period. Other interventions that may affect psychological status, such as cognitive behavioral therapy, will not be prohibited; however, their content will be recorded and considered in the interpretation of the findings. If antidepressants are prescribed by the treating physician, information regarding their use, indication, and medication names will be recorded. Interventions specifically intended to modify attentional bias, including ABM and tasks identical or similar to the study tasks, will not be implemented during the study period. If such interventions are delivered for unavoidable clinical reasons, their content and timing will be documented and considered in the analysis or interpretation. An overview of the assessment schedule is presented in Fig 3.

**Fig 3.**
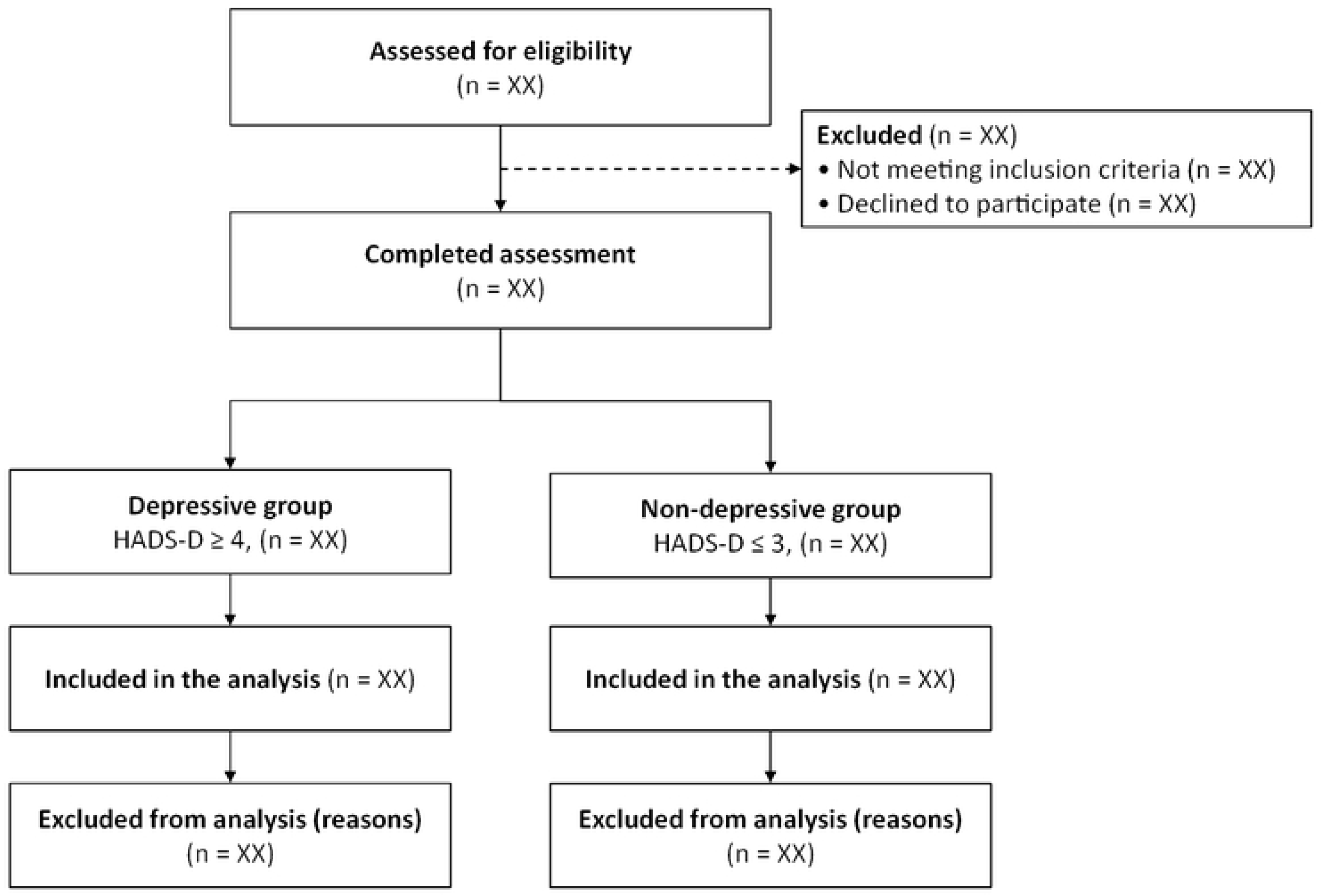
Flowchart of study participants. Potential participants will be screened for eligibility. Patients who do not meet the inclusion criteria, meet the exclusion criteria, or decline participation will be excluded. Eligible participants who provide written informed consent will undergo study assessments and will be classified into depressive symptom and non-depressive symptom groups according to their Hospital Anxiety and Depression Scale–Depression (HADS-D) score for predefined analyses.

### Measures

Regarding evaluation scales that were originally designed for self-administration (HADS and EuroQol 5 Dimensions 5 Levels [EQ-5D-5L]), because the target population is expected to consist predominantly of older patients, the examiner will read the questions aloud and record the responses using an interviewer-administered format. Previous research evaluating brief screening instruments, such as the Geriatric Depression Scale and the MMSE, in older populations demonstrated that face-to-face interview methods resulted in fewer missing values and yielded higher data quality compared with self-administered mail surveys (Smeeth, 2001).

Similarly, studies evaluating the EQ-5D indicate that older respondents frequently require assistance, making interviewer-administered surveys effective (Marten, 2021). Furthermore, the developer of the instrument, the EuroQol Group, officially approves interviewer-administered formats for respondents with reading or writing difficulties, explicitly stating that data obtained via interviewer administration can be used interchangeably with the self-completed version.

Based on these findings, modifying the administration of these questionnaires to an oral readout format in this study was judged to be methodologically appropriate.

### Depressive symptoms

Depressive symptoms will be assessed using the depression subscale of the Hospital Anxiety and Depression Scale (HADS-D) [25]. The Japanese version translated by Kitamura will be used [26]. The HADS is a 14-item self-report measure consisting of seven anxiety items and seven depression items and is designed to assess symptoms experienced during the previous week. Therefore, this study will include patients at least 7 days after stroke onset. Each item is scored on a 4-point scale ranging from 0 to 3, and the scale imposes a relatively low burden on patients [27]. The HADS-D total score will be treated primarily as a continuous variable. In addition, based on a recommended cutoff for stroke populations [28], participants with a HADS-D score of 4 or higher will be classified as having depressive symptoms for predefined group-based analyses.

### Anxiety symptoms

Anxiety symptoms will be assessed using the anxiety subscale of the Hospital Anxiety and Depression Scale (HADS-A) [27]. HADS-A scores will be used as an additional psychological variable and as a potential confounder in multivariable analyses.

### Cognitive function

Cognitive function will be assessed using the MMSE. In addition to serving as an eligibility criterion, MMSE scores will be analyzed as a continuous variable and will also be used to classify participants according to cognitive status using a predefined cutoff based on prior literature [29]. This study includes participants with an MMSE score ≥ 24, and a cutoff value of 27/28 points will be used in the statistical analysis to differentiate between cognitively healthy individuals and those with MCI. Cognitive function will be examined as a potential moderator of the association between depressive symptoms and attentional bias.

### ADL and quality of life

ADL will be assessed using the Functional Independence Measure (FIM). Health-related quality of life will be assessed using the Japanese version of the EQ-5D-5L. These variables will be used to characterize the sample and to explore their associations with depressive symptoms and attentional bias.

### Clinical and demographic variables and assessment of handicap

The following clinical and demographic variables will be collected from medical records and participant interviews: lesion location; level of handicap assessed using the modified Rankin Scale (mRS); age; sex; handedness; affected side; days since stroke onset; medical history; and educational history. These variables will be used to characterize the sample and, where appropriate, as covariates in statistical analyses. Baseline demographic and clinical characteristics will be summarized in Table 1. Cognitive, emotional, quality-of-life, and functional measures will be summarized in Table 2.

**Table 1.**
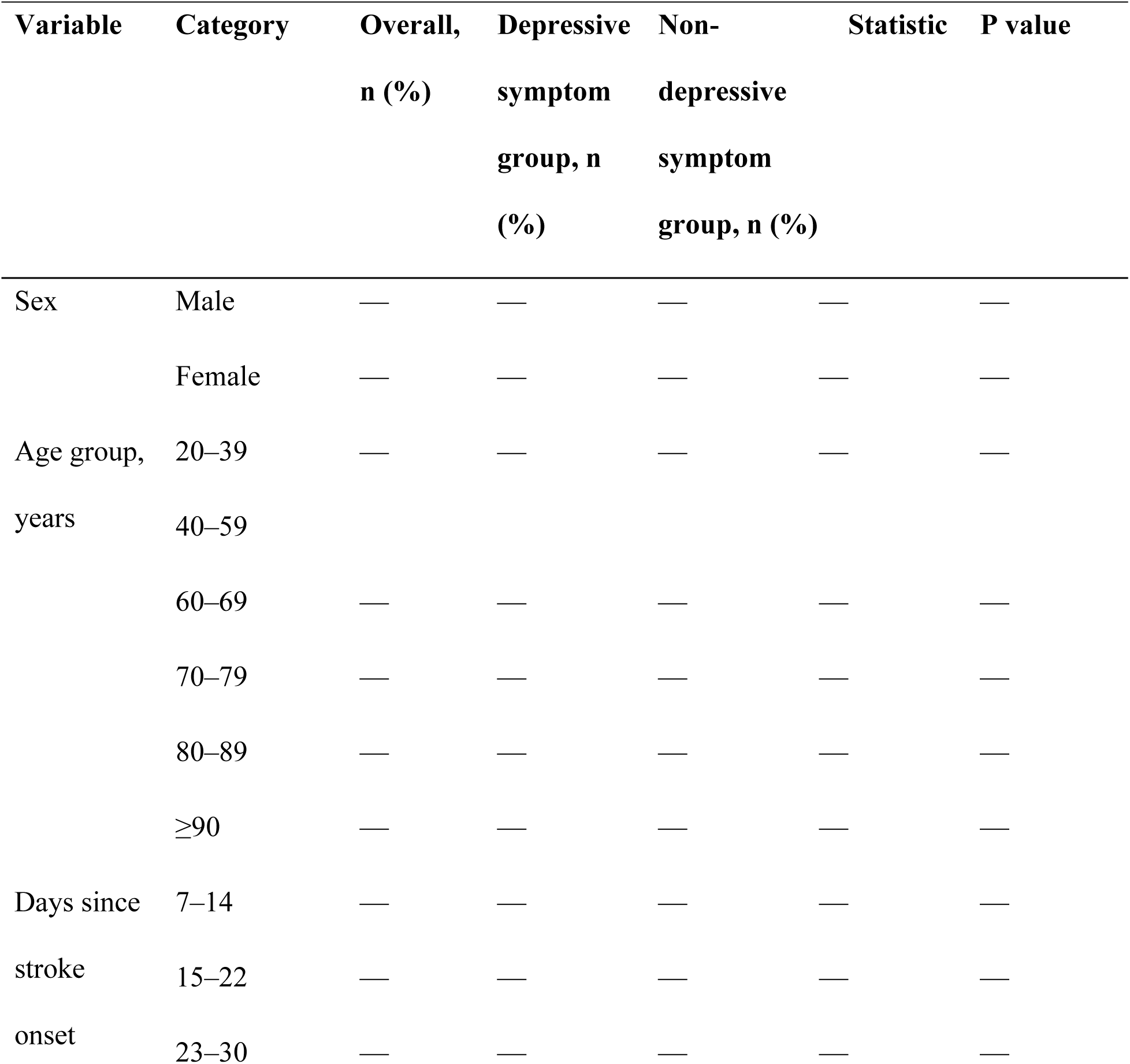
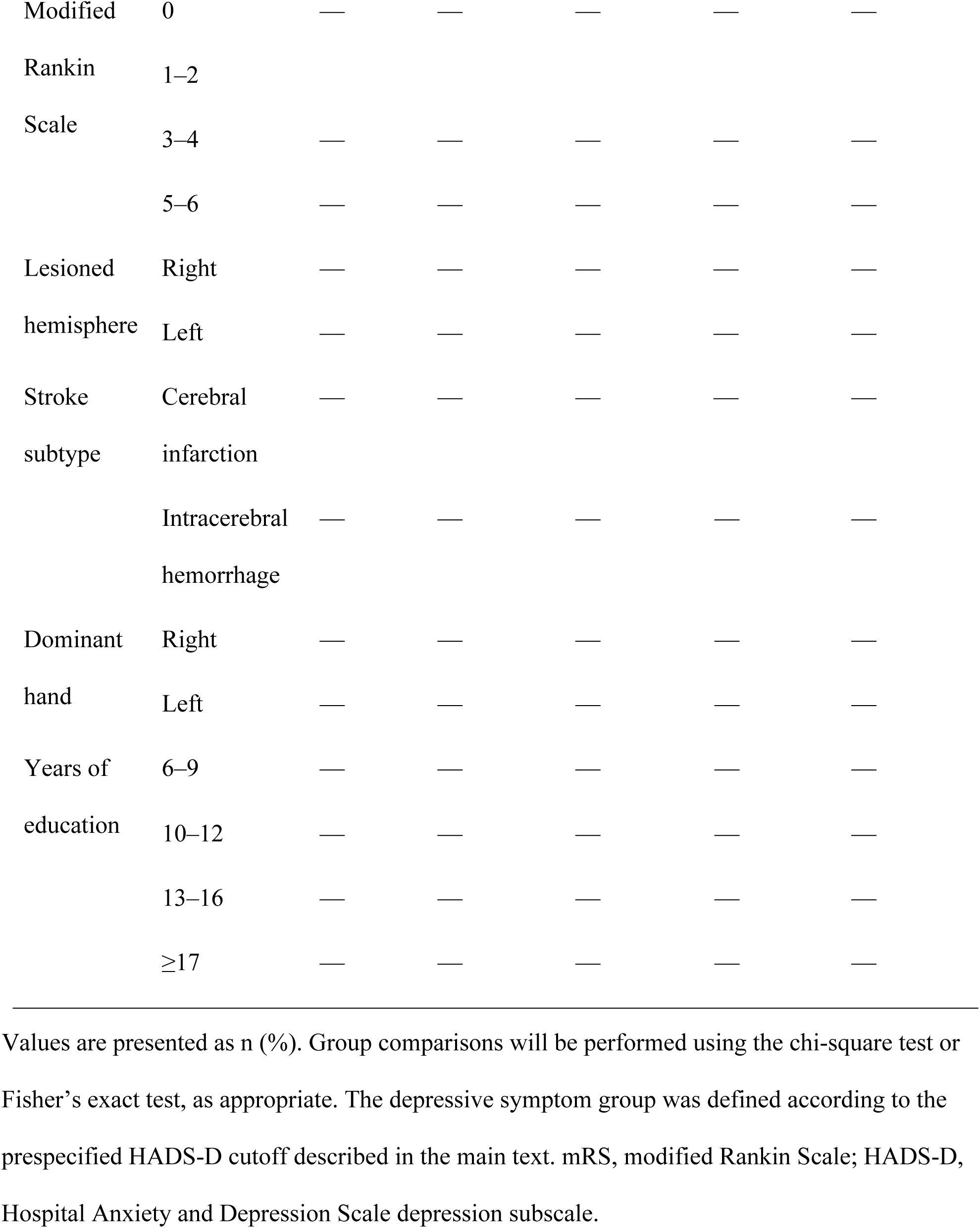
Demographic and clinical characteristics of the participants.

**Table 2.**
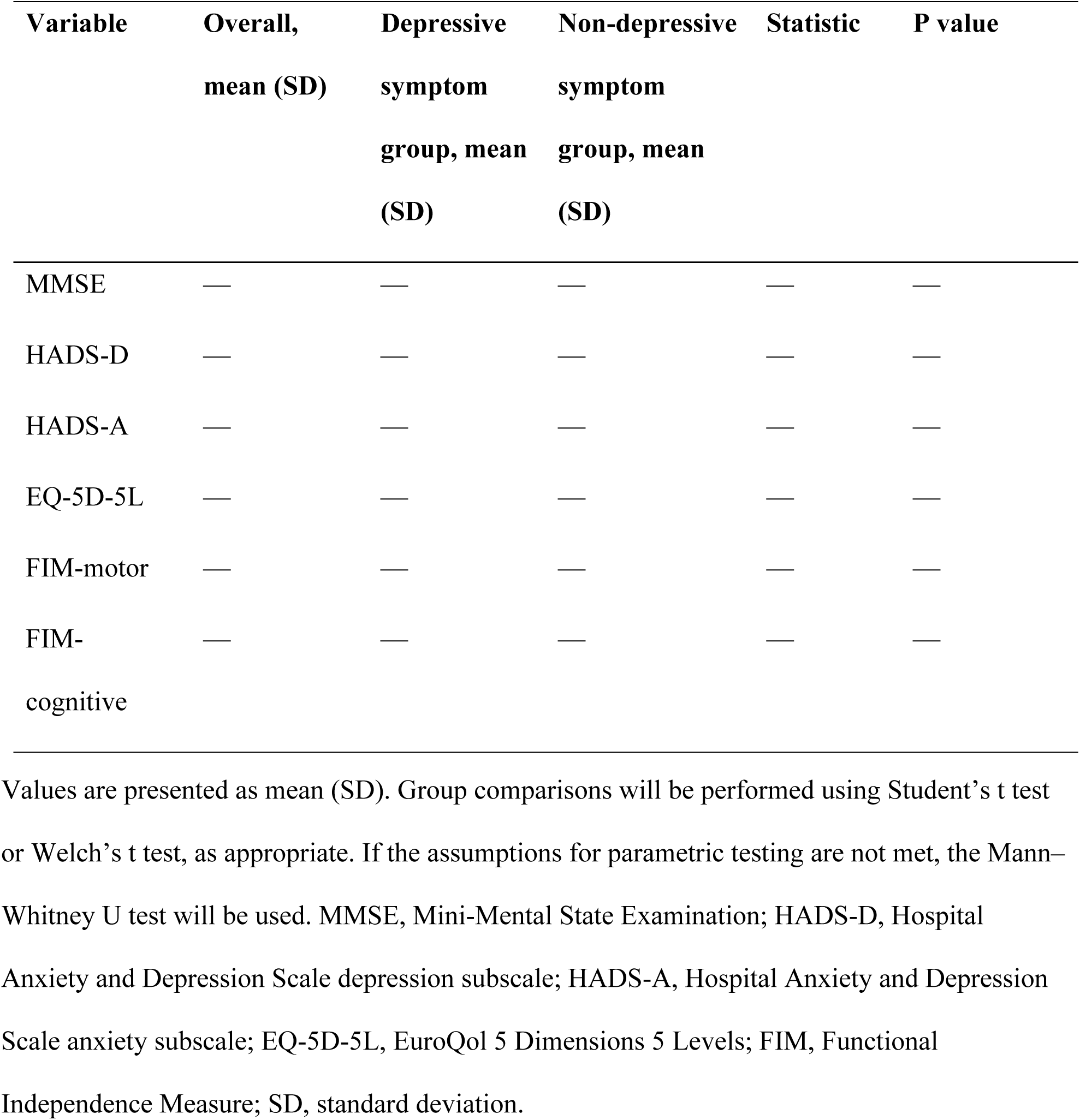
Comparison of psychological, cognitive, quality-of-life, and functional measures between the depressive symptom and non-depressive symptom groups.

### Attentional bias assessment

Attentional bias will be assessed using the evaluation mode of the ABM online training software (ABOT; Saitama Prefectural University, 2022). Four task conditions will be administered: DPT-Face, DPT-Word, CTT-Face, and CTT-Word. The experimental procedure and scoring concept for the DPT are shown in Fig 4. The experimental procedure and scoring concept for the CTT are shown in Fig 5. Attentional bias scores will be calculated separately for each condition. More negative scores will indicate a stronger bias toward negative stimuli, operationalized as either selective attention to aversive stimuli or difficulty disengaging attention from aversive stimuli, depending on the task structure.

**Fig 4.**
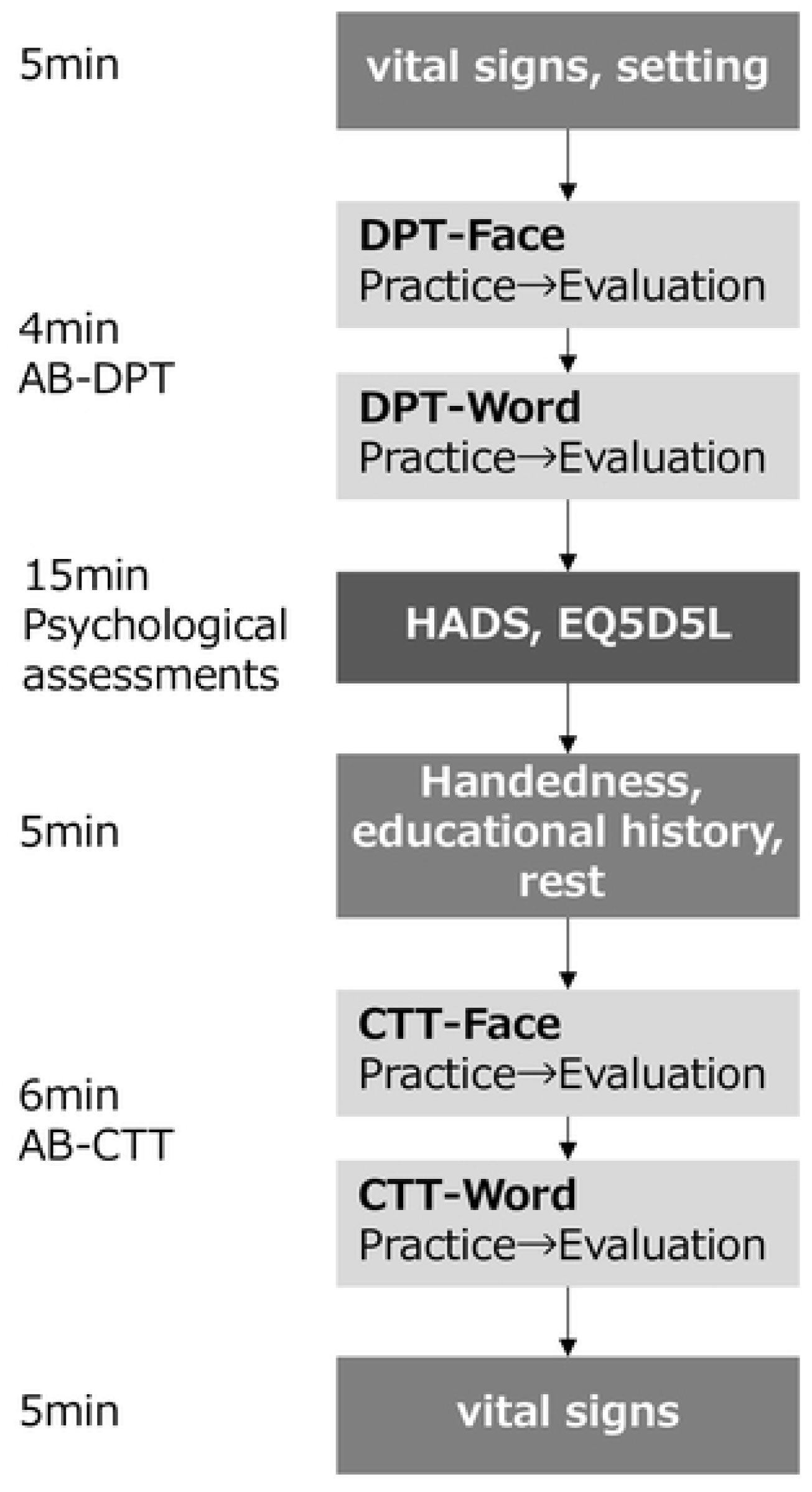
Study assessment schedule. The planned assessment lasts approximately 40 minutes and includes preparation (vital sign check, postural adjustment, and task explanation), practice and assessment for the DPT facial and word tasks, psychological assessments, collection of handedness and educational history with rest provided as needed, practice and assessment for the CTT facial and word tasks, and a post-assessment vital sign check. If necessary, the assessment may be postponed depending on the participant’s physical condition.

**Fig 5.**
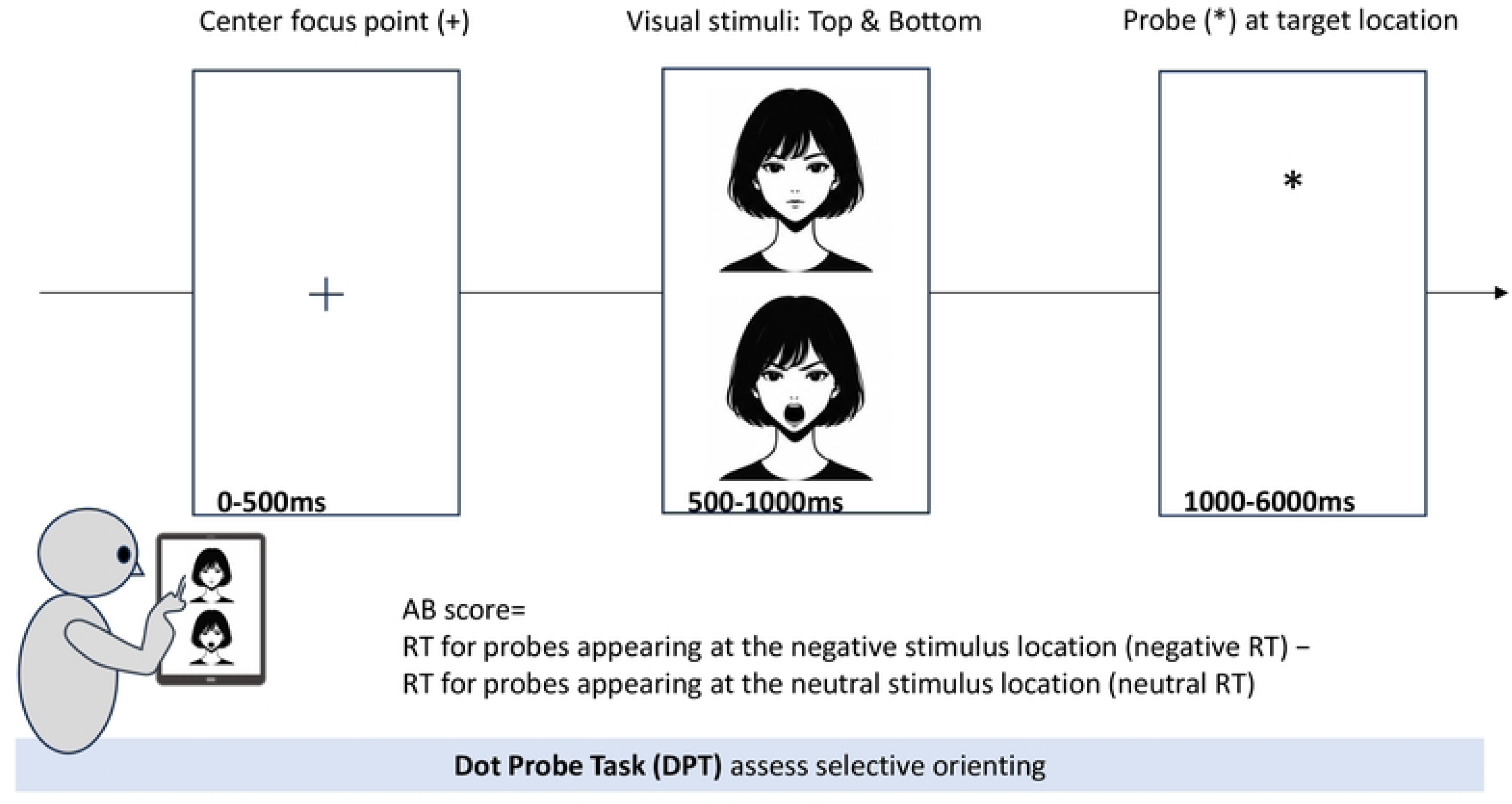
Dot-probe task (DPT). A fixation point is presented at the center of the screen for 500 ms, followed by the simultaneous presentation of two vertically aligned stimuli (an aversive stimulus and a neutral stimulus) for 500 ms. A probe then appears in the location of either the aversive or the neutral stimulus, and the participant is instructed to respond as quickly as possible. Reaction time from probe onset to response is recorded. In the assessment condition, probes appear equally often in the aversive and neutral stimulus locations. The attentional bias score is calculated by subtracting the reaction time for probes replacing neutral stimuli from the reaction time for probes replacing aversive stimuli. More negative scores indicate stronger selective attention toward aversive stimuli. In the DPT, the cue stimuli are presented for 500 ms, and the probe is presented immediately after cue offset without an interstimulus interval (ISI), resulting in a stimulus-onset asynchrony (SOA) of 500 ms.

In the DPT, a fixation point will be presented at the center of the screen for 500 ms, followed by the simultaneous presentation of two stimuli for 500 ms in vertically aligned positions. One aversive stimulus and one neutral stimulus will be presented, and a probe will then appear in the location of either the aversive or neutral stimulus. Participants will be instructed to respond to the probe as quickly and accurately as possible. For DPT-Face and DPT-Word, attentional bias scores will be calculated by subtracting the reaction time for probes replacing neutral stimuli from the reaction time for probes replacing aversive stimuli. More negative values will indicate faster responses to probes in the aversive stimulus location and, therefore, stronger selective attention toward aversive stimuli [18].

In the CTT, an emotional stimulus will be presented unilaterally on the left or right side of the screen for 1500 ms, followed by a probe that appears either on the same side as the emotional cue or on the opposite side. For CTT-Face and CTT-Word, attentional bias scores will be calculated by subtracting the reaction time for probes appearing opposite the aversive cue from the reaction time for probes appearing on the same side as the aversive cue. More negative values will indicate greater difficulty disengaging attention from aversive stimuli [19].

Facial stimuli consisted of aversive (angry) and neutral facial expressions presented in pairs. The stimuli were selected from the Japanese Female Facial Expression (JAFFE) database, which consists of images of Japanese women displaying six basic facial expressions—happiness, sadness, fear, anger, surprise, and disgust—as well as neutral expressions [30]. Emotional ratings by Japanese evaluators have been conducted for these images, and the correspondence between image features extracted using Gabor filter–based methods and human emotional ratings has been examined [30]. Previous studies have reported that JAFFE expressions represent naturally elicited facial expressions within a Japanese cultural context and that Japanese participants demonstrate relatively high recognition accuracy for JAFFE expressions [31]. In the present study, eight aversive (angry) facial expressions and eight neutral facial expressions (16 images in total) were selected from the JAFFE database. These eight pairs of facial stimuli will be presented in random order for a total of 128 trials. Examples of the facial stimuli used in this study are shown in Fig 6.

**Fig 6.**
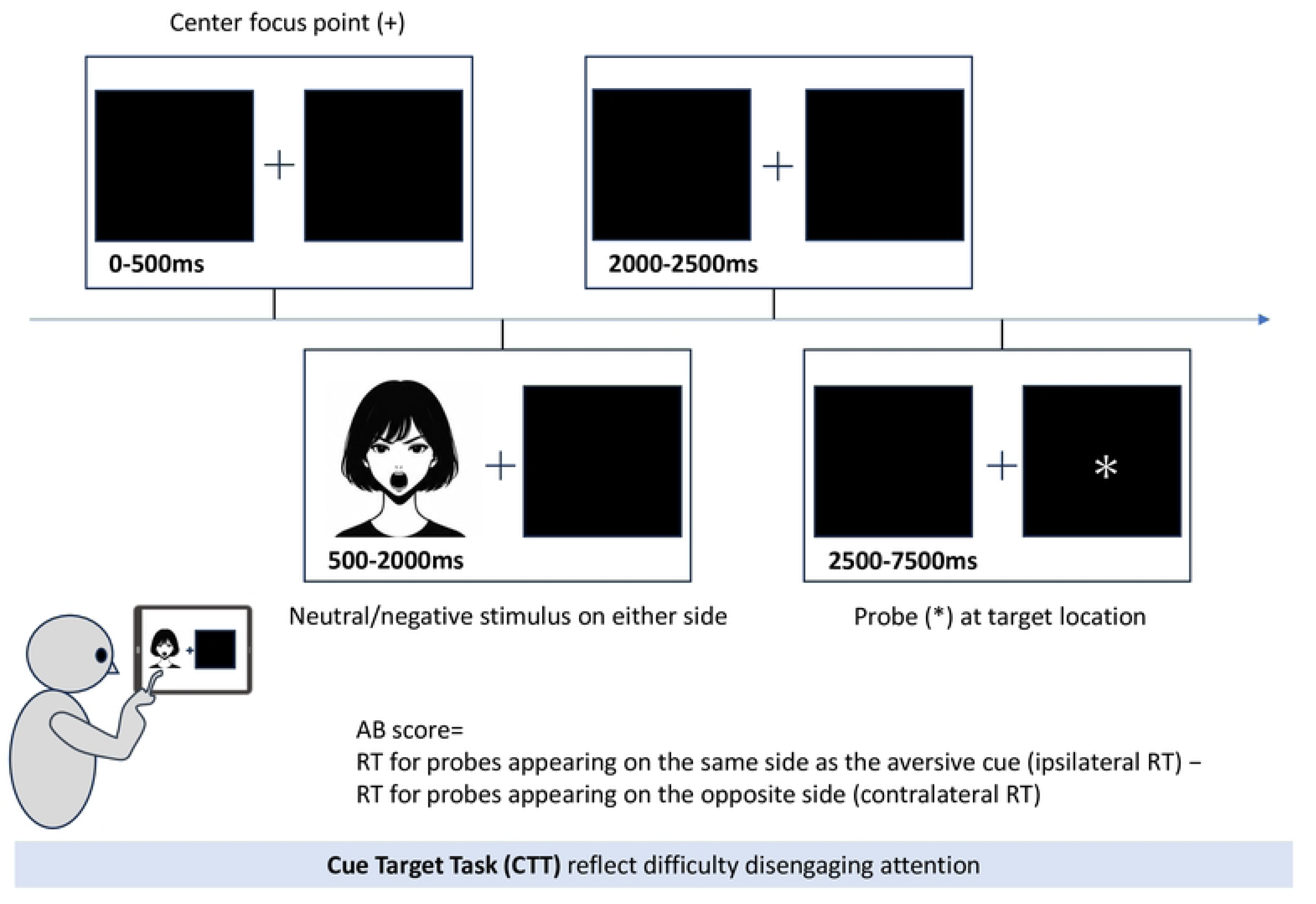
Cue–target task (CTT). An emotional stimulus is presented on either the left or right side of the screen for 1500 ms, followed by a probe that appears either on the same side or on the opposite side. Reaction times for probes appearing on the same side as the aversive cue and on the opposite side are recorded. The attentional bias score is calculated by subtracting the reaction time for probes appearing opposite the aversive cue from the reaction time for probes appearing on the same side as the aversive cue. More negative scores indicate greater difficulty disengaging attention from aversive stimuli. In the CTT, the cue stimulus is presented for 1500 ms, followed by a 500-ms interstimulus interval (ISI) before probe presentation, resulting in a stimulus-onset asynchrony (SOA) of 2000 ms.

**Fig 7.**
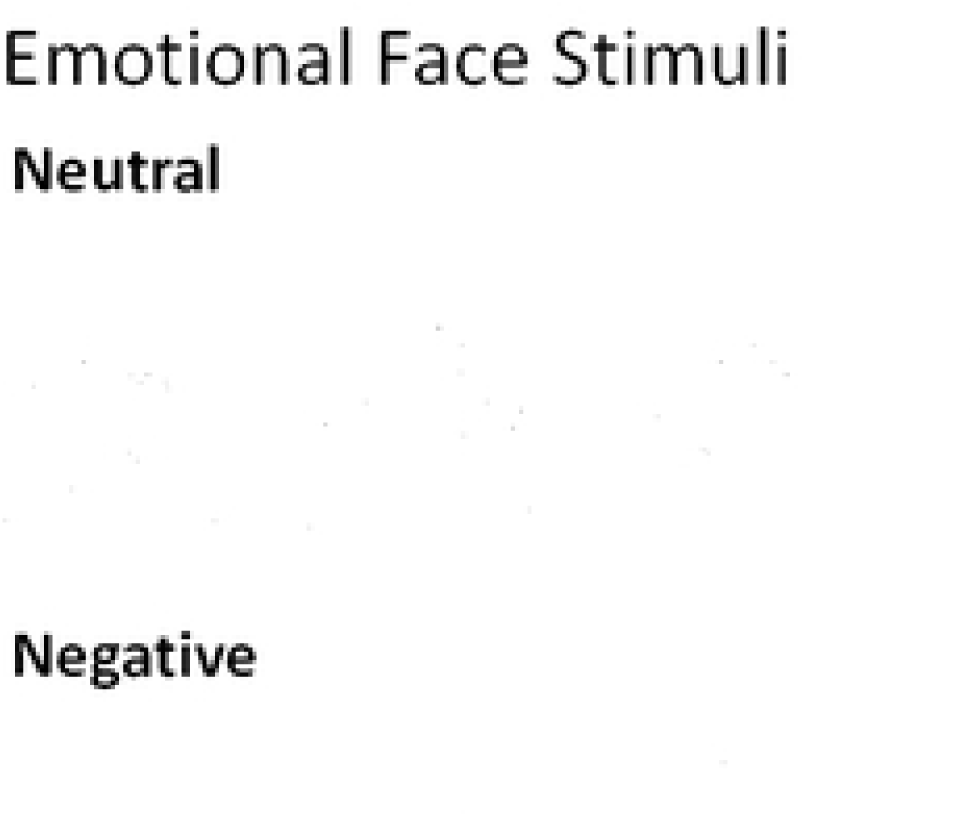
Examples of emotional facial stimuli. Facial stimuli used in the task consisted of aversive (angry) and neutral facial expressions selected from the Japanese Female Facial Expression (JAFFE) database. Eight aversive–neutral pairs (16 images in total) were used and will be presented repeatedly in randomized order during the task.

The word stimuli were selected from a set of words whose emotional valence had been evaluated in a previous study [32]. Emotional valence was rated on a 7-point scale, with lower scores indicating more positive words and higher scores indicating more negative words. The specific word stimuli used in this study include neutral words (valence: 3.49–3.80) including humanity, preparation, muscle, connection, basic, daytime, consent, and choice, and aversive words (valence: 6.03–6.49) including discrimination, tragedy, banishment, despair, prejudice, anxiety, worst, and disease. Examples of the word stimuli are shown in Table 3.

**Table 3.**
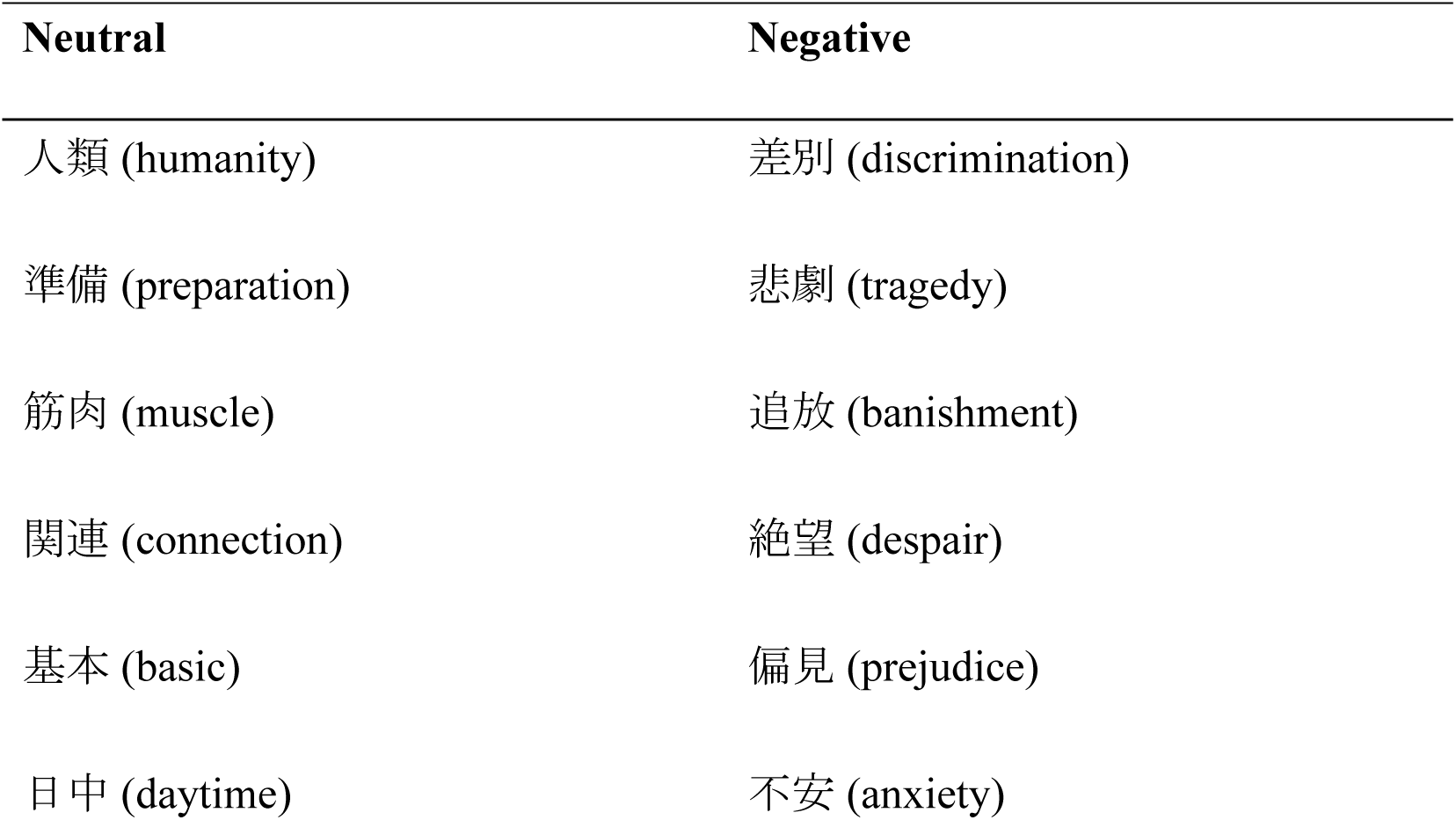
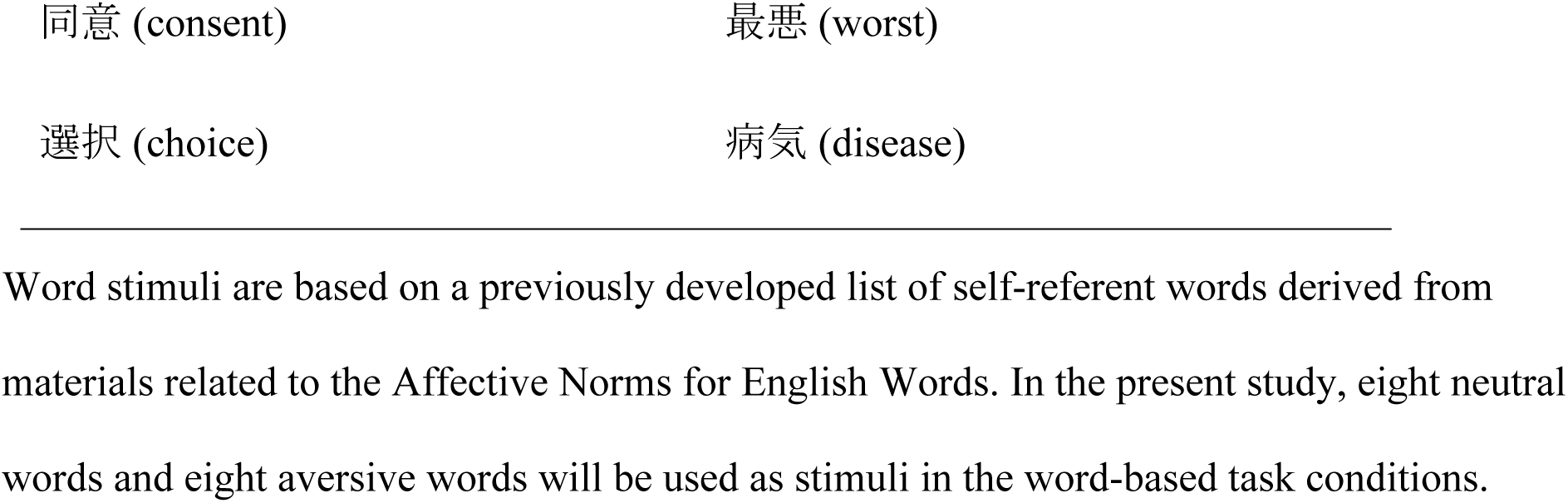
Word stimuli used in the attentional bias tasks Neutral Negative.

| Neutral | Negative |
| --- | --- |
| 人類 (humanity) | 差別 (discrimination) |
| 準備 (preparation) | 悲劇 (tragedy) |
| 筋肉 (muscle) | 追放 (banishment) |
| 関連 (connection) | 絶望 (despair) |
| 基本 (basic) | 偏見 (prejudice) |
| 日中 (daytime) | 不安 (anxiety) |

Word stimuli are based on a previously developed list of self-referent words derived from materials related to the Affective Norms for English Words. In the present study, eight neutral words and eight aversive words will be used as stimuli in the word-based task conditions.

### Variables

The primary explanatory variable will be depressive symptom severity, measured using the continuous HADS-D score. In predefined group-based analyses, participants will also be classified into a depressive symptom group and a non-depressive symptom group using a HADS-D cutoff score of 4 [28]. The primary outcome variables will be the four attentional bias indices obtained from the ABOT evaluation mode: DPT-Face, DPT-Word, CTT-Face, and CTT-Word. Secondary variables will include anxiety symptoms, cognitive function, ADL, health-related quality of life, lesion location, level of handicap, age, sex, handedness, affected side, days since stroke onset, medical history, and educational history. Age, sex, cognitive function, anxiety symptoms, days since stroke onset, antidepressant use, concomitant psychological interventions, lesion location, and level of handicap will be considered potential confounders, as appropriate. Cognitive function will also be examined as a potential effect modifier.

### Bias control

Several measures will be implemented to address potential threats to the validity of this study. To minimize selection bias, patients who meet the eligibility criteria will be identified as comprehensively as possible at each participating site within the limits of routine clinical practice, thereby reducing arbitrariness in participant selection. To reduce information bias, the assessment setting (private room or bedside) and the participant’s physical condition on the day of assessment will be recorded to account for environmental influences. To ensure the validity of the attentional bias indices, participants who do not achieve the predefined accuracy threshold during the practice task (at least 70% accuracy in the first 10 trials) will be excluded from the main analyses, and the number of such exclusions and the reasons for exclusion will be documented. In addition, major potential confounding factors, including anxiety symptoms, cognitive function, age, and sex, will be included as covariates in the statistical analyses to allow for appropriate adjustment.

### Sample size

No previous study has reported attentional bias scores in patients with acute stroke. Therefore, the required sample size for the present study was estimated using G*Power based on an assumed effect size of 0.8, an alpha level of 0.05, and statistical power of 0.80. The ratio between participants with and without depressive symptoms was set at 1:3 based on the reported frequency of depressive symptoms after stroke [2]. This calculation yielded a required sample of 17 participants in the depressive symptom group and 51 participants in the non-depressive symptom group, for a total of 68 participants. To account for dropout and missing data, the target sample size was increased by 10%, resulting in a final target of 76 participants, including 19 participants with depressive symptoms and 57 without depressive symptoms. Because the present study is exploratory and prior data are limited, the effect size estimates obtained from this study are expected to serve as pilot data for future confirmatory research.

### Data collection and management

After written informed consent is obtained, participants will be registered in the study. Depressive symptoms, anxiety symptoms, cognitive function, attentional bias, and health-related quality of life will be assessed by study investigators during routine clinical hours. ADL will be evaluated by the study investigators using information from treating therapists, medical records, and routine rehabilitation assessments. Clinical variables, including age, sex, educational history, medical history, days since stroke onset, lesion location, and level of handicap, will be collected from medical records.

Participants may withdraw their consent to participate in the study at any time. Personally identifiable information will be managed only at each participating institution. After deidentification, all study data will be stored on security-protected devices under the control of the study investigators. Access to identifiable information and study data will be restricted to authorized research personnel only.

### Statistical analysis

For the primary analyses, participants will be classified into depressive symptom and non-depressive symptom groups based on a predefined HADS-D cutoff score. Additional exploratory analyses will examine HADS-D scores as continuous variables. For the attentional bias assessment, trials with reaction times (RTs) shorter than 300 ms or longer than 3,000 ms [33], as well as trials with incorrect responses (responses opposite to the probe location), will be excluded prior to statistical analysis.

The primary hypothesis is that the direction and magnitude of attentional bias differ between participants with and without depressive symptoms. For each of the four attentional bias scores, descriptive statistics, including means, standard deviations, and 95% confidence intervals, will be calculated. The distribution of each attentional bias score will be examined using the Shapiro–Wilk test. To determine whether attentional bias is present within each group, attentional bias scores will be compared with zero using a one-sample t test for normally distributed data or the Wilcoxon signed-rank test for non-normally distributed data. Positive values will be interpreted as indicating a bias toward aversive stimuli, and negative values as indicating a bias away from aversive stimuli. Between-group comparisons will be performed using the independent-samples t test for normally distributed data or the Mann–Whitney U test for non-normally distributed data. Effect sizes will be calculated as Cohen’s d for t tests and r for Mann–Whitney U tests. Within-group comparisons of attentional bias scores against zero will be summarized in Table 4. Between-group comparisons of attentional bias scores will be summarized in Table 5.

**Table 4.** Within-group comparison of attentional bias scores against zero.

| Group | Condition | Mean (SD) | 95% CI | Statistic | P value | Effect size |
| --- | --- | --- | --- | --- | --- | --- |
| Depressive symptom group | DPT-Face | — | — | — | — | — |
|  | DPT-Word | — | — | — | — | — |
|  | CTT-Face | — | — | — | — | — |
|  | CTT-Word | — | — | — | — | — |
| Non-depressive symptom group | DPT-Face | — | — | — | — | — |
|  | DPT-Word | — | — | — | — | — |
|  | CTT-Face | — | — | — | — | — |
|  | CTT-Word | — | — | — | — | — |

Values are presented as mean (SD). Attentional bias scores will be compared with zero within each group using a one-sample t test or the Wilcoxon signed-rank test, as appropriate. Negative values indicate stronger bias toward aversive stimuli according to the task-specific scoring definitions described in the text. Abbreviations: DPT, dot-probe task; CTT, cue–target task; CI, confidence interval; SD, standard deviation.

**Table 5.** Comparison of attentional bias scores between the depressive symptom and non-depressive symptom groups.

| Condition | Depressive<br>symptom<br>group,<br>mean (SD) | Non-depressive<br>symptom<br>group,<br>mean (SD) | 95% CI<br>for<br>difference | Statistic | P value | Effect size |
| --- | --- | --- | --- | --- | --- | --- |
| DPT-Face | — | — | — | — | — | — |
| DPT-Word | — | — | — | — | — | — |
| CTT-Face | — | — | — | — | — | — |
| CTT-Word | — | — | — | — | — | — |

Values are presented as mean (SD). Group comparisons will be performed using Student's t test, Welch's t test, or the Mann–Whitney U test, as appropriate. Negative values indicate stronger bias toward aversive stimuli according to the task-specific scoring definitions described in the text. Abbreviations: DPT, dot-probe task; CTT, cue–target task; CI, confidence interval; SD, standard deviation.

Secondary analyses will examine the effects of stimulus type and task format on attentional bias. In participants with depressive symptoms, a two-way repeated-measures analysis of variance (ANOVA) will be conducted with stimulus type (facial vs word) and task type (dot-probe vs cue–target) as within-subject factors to examine differences in attentional bias magnitude. If a significant interaction is observed, post hoc comparisons of simple main effects will be conducted using paired t tests.

In all participants, a three-way mixed ANOVA will be conducted with group (depressive symptom vs non-depressive symptom) as the between-subject factor and stimulus type (facial vs word) and task type (dot-probe vs cue–target) as within-subject factors. Main effects, two-way interactions, and the three-way interaction will be examined. If significant interactions are detected, simple main-effect analyses will be performed. Results of the two-way repeated-measures ANOVA in the depressive symptom group will be summarized in Table 6. Results of the three-way mixed ANOVA across all participants will be summarized in Table 7. Simple main-effect analyses, when conducted, will be summarized in Table 8.

**Table 6.**
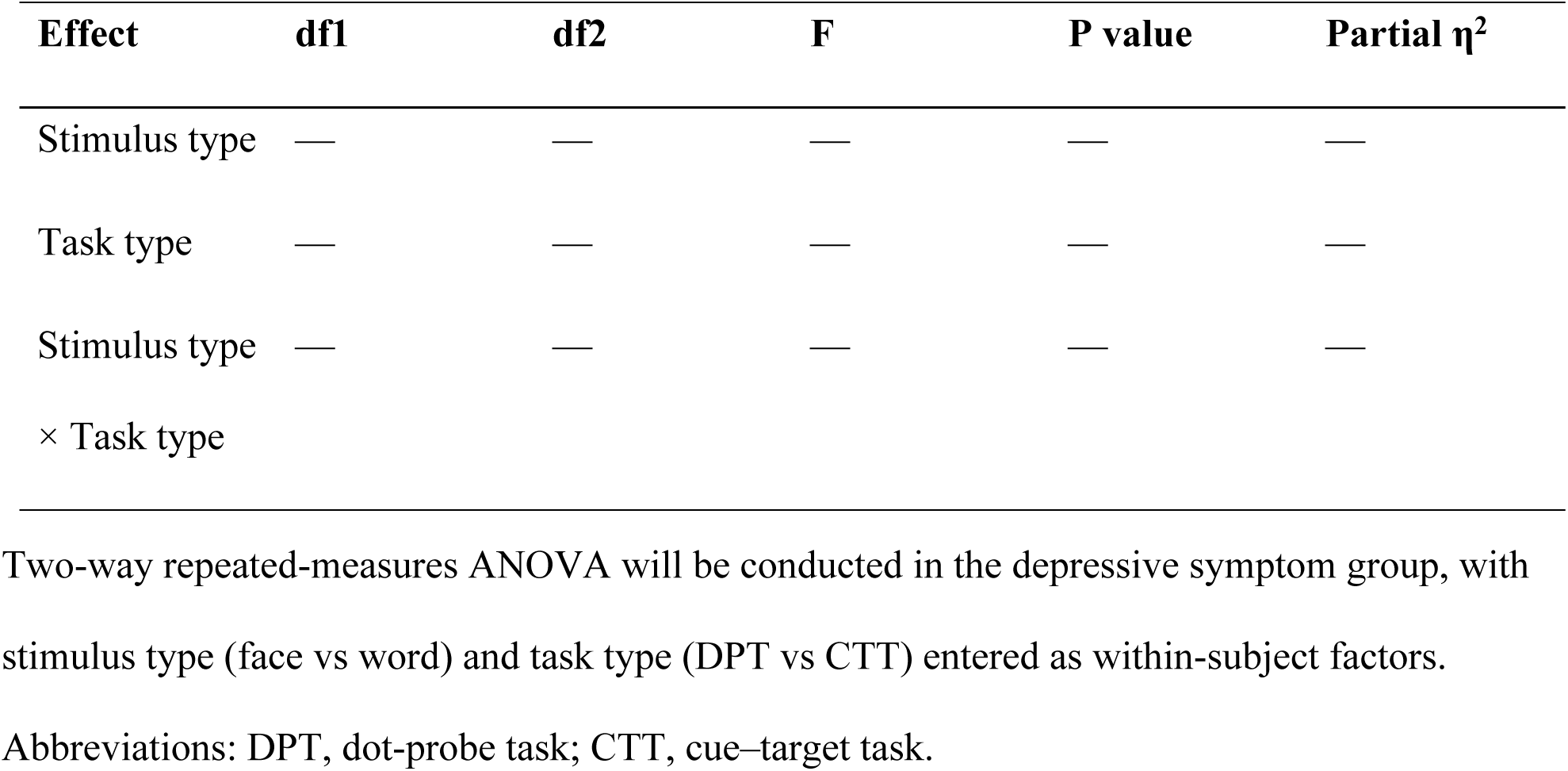
Effects of stimulus type and task type on attentional bias scores in the depressive symptom group: two-way repeated-measures ANOVA.

**Table 7.**
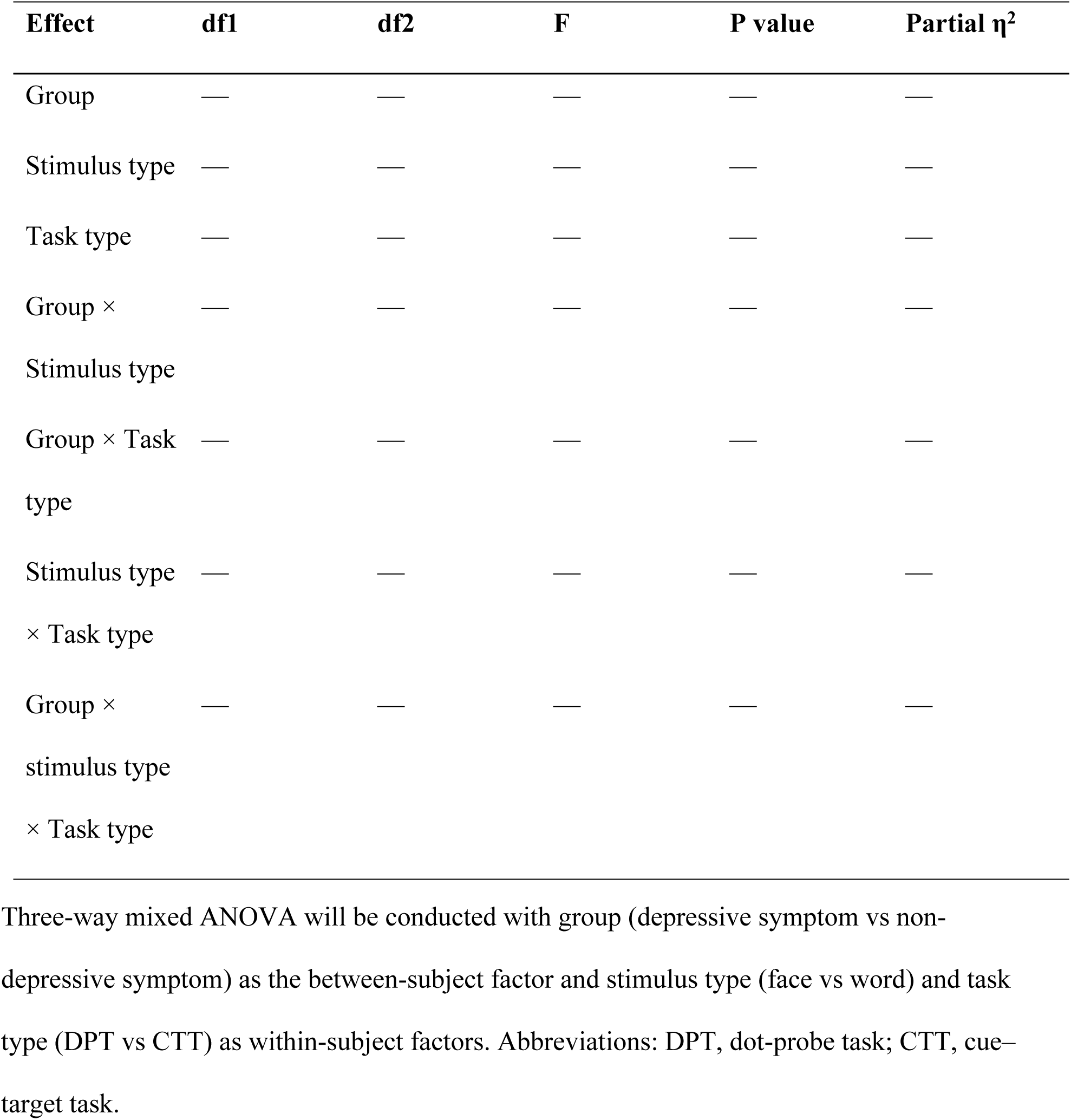
Effects of group, stimulus type, and task type on attentional bias scores: three-way mixed ANOVA.

| Effect | df1 | df2 | F | P value | Partial $\eta^2$ |
| --- | --- | --- | --- | --- | --- |
| Group | — | — | — | — | — |
| Stimulus type | — | — | — | — | — |
| Task type | — | — | — | — | — |
| Group × Stimulus type | — | — | — | — | — |
| Group × Task type | — | — | — | — | — |
| Stimulus type × Task type | — | — | — | — | — |
| Group × stimulus type × Task type | — | — | — | — | — |

Three-way mixed ANOVA will be conducted with group (depressive symptom vs non-depressive symptom) as the between-subject factor and stimulus type (face vs word) and task type (DPT vs CTT) as within-subject factors. Abbreviations: DPT, dot-probe task; CTT, cue-target task.

**Table 8.** Results of simple main-effect analyses using paired t tests.

| Group | Stimulus type | Comparison | Mean difference | 95% CI | Statistic | P value | Effect size |
| --- | --- | --- | --- | --- | --- | --- | --- |
| Depressive symptom | Face | DPT vs CTT | — | — | — | — | — |
| Depressive symptom | Word | DPT vs CTT | — | — | — | — | — |
| Non-depressive symptom | Face | DPT vs CTT | — | — | — | — | — |
| Non-depressive symptom | Word | DPT vs CTT | — | — | — | — | — |

Simple main effects will be examined using paired t tests. Comparisons will be conducted according to the significant interaction effects identified in the ANOVA models. Abbreviations: DPT, dot-probe task; CTT, cue–target task; CI, confidence interval.

Additional analyses will examine the continuous association between depressive symptom severity and attentional bias. Spearman’s rank correlation coefficients will be calculated to assess the associations between continuous HADS-D scores and each attentional bias index. Multivariable regression analyses will then be conducted with each attentional bias score as the dependent variable and the HADS-D score as the independent variable, adjusting for age, sex, MMSE score, days since stroke onset, and HADS-A score. To explore whether cognitive function modifies the relationship between depressive symptoms and attentional bias, an interaction term (MMSE score × HADS-D score) will be entered into the regression models. Associations between depressive symptom severity and attentional bias scores will be summarized in Table 9. Multiple regression analyses examining depressive symptoms and covariates as predictors of attentional bias scores will be summarized in Table 10.

**Table 9.**
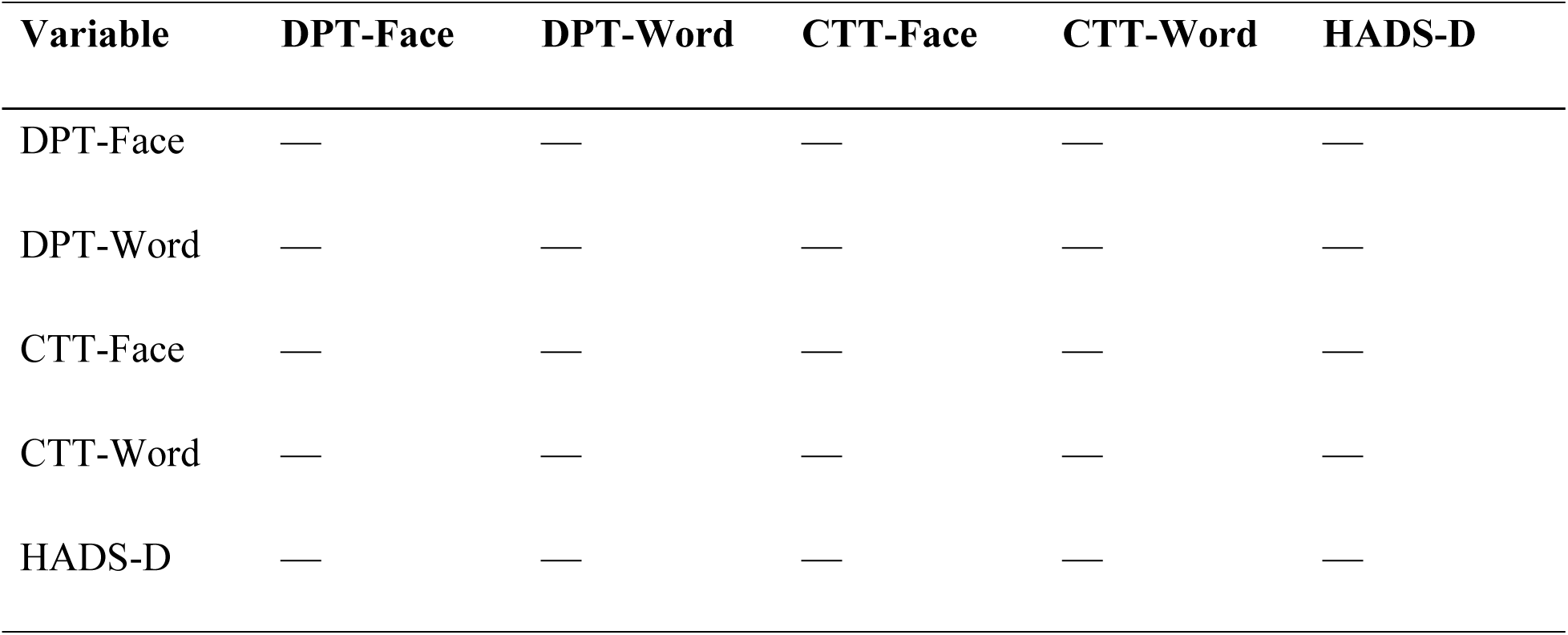
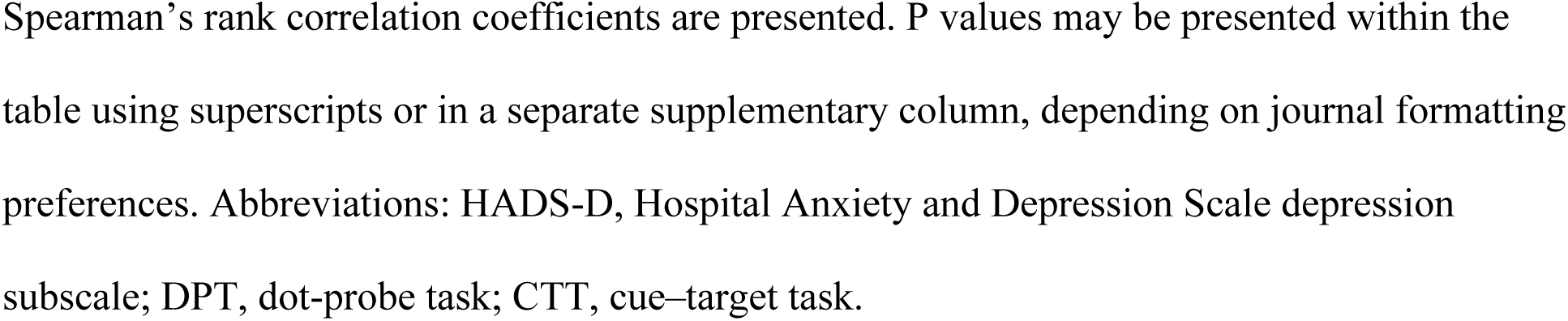
Correlations between HADS-D scores and attentional bias scores.

| Variable | DPT-Face | DPT-Word | CTT-Face | CTT-Word | HADS-D |
| --- | --- | --- | --- | --- | --- |
| DPT-Face | — | — | — | — | — |
| DPT-Word | — | — | — | — | — |
| CTT-Face | — | — | — | — | — |
| CTT-Word | — | — | — | — | — |
| HADS-D | — | — | — | — | — |

Spearman's rank correlation coefficients are presented. P values may be presented within the table using superscripts or in a separate supplementary column, depending on journal formatting preferences. Abbreviations: HADS-D, Hospital Anxiety and Depression Scale depression subscale; DPT, dot-probe task; CTT, cue–target task.

**Table 10.** Planned multivariable regression analyses of depressive symptoms and covariates as predictors of attentional bias scores.

| Outcome | $\beta$ for HADS-D | 95% CI | P value | $\beta$ for HADS-D × MMSE | P value for interaction |
| --- | --- | --- | --- | --- | --- |
| DPT-Face | — | — | — | — | — |
| DPT-Word | — | — | — | — | — |
| CTT-Face | — | — | — | — | — |
| CTT-Word | — | — | — | — | — |

The number of participants with missing data for each variable will be documented. Analyses will be conducted using the available data for each outcome. Participants who are unable to complete a specific attentional bias task or who do not meet the predefined practice accuracy threshold will be excluded from analyses involving that task, and the number of such exclusions and the reasons for exclusion will be reported transparently. Because the study is exploratory, interpretation of the findings will take into account the influence of multiple testing and potential protocol-related variability. Supplementary analyses may be conducted, as appropriate, to examine the robustness of the findings after excluding participants with protocol deviations or clinically relevant factors that may substantially affect task performance or interpretation.

All statistical analyses will be performed using jamovi (version 2.6.23) and R (version 4.3 or later). Statistical significance will be set at p < 0.05. In interpreting the findings, emphasis will be placed not only on p values but also on effect sizes and 95% confidence intervals.

### Ethics approval, registration, and study status

This study has been registered with the UMIN Clinical Trials Registry (UMIN000059166). At the time of manuscript submission, ethical approval has been obtained from the ethics committees of the investigators’ affiliated university (No. 25159), Niiza Shiki Central General Hospital (No. 2025-5), Totsuka Kyoritsu Izumino Hospital (approval number not assigned), and Nishitokyo Chuo General Hospital (No. 2026-2). Written informed consent will be obtained from all participants before enrollment. The study will be conducted in accordance with the principles of the Declaration of Helsinki.

Participant recruitment will begin after ethics approval has been obtained at each participating institution. Recruitment is scheduled from Jun 10, 2026, to November 30, 2026. Because this is a cross-sectional study, each participant will complete the study after a single assessment session, and no follow-up will be performed. At the protocol stage, no study results are available.

## Discussion

This study protocol describes a multicenter cross-sectional observational study designed to examine the association between depressive symptoms and attentional bias to emotional stimuli in patients with acute stroke. PSD is common in the early phase after stroke [2,5]; however, the characteristics of attentional bias during this stage have not been sufficiently characterized. It remains unclear whether attentional bias is associated with depressive symptoms in patients with acute stroke and whether such bias can provide a clinically meaningful basis for identifying candidates for future attentional bias modification (ABM) interventions. Previous work by our group in patients in the convalescent stage after stroke found that attentional orienting was related to cognitive function rather than depressive symptoms, with depressed patients without MCI orienting more rapidly toward neutral rather than aversive faces [13]. The present study is therefore expected to provide foundational evidence for the future development of ABM-based interventions in the acute stage after stroke.

A key strength of this protocol is its multidimensional assessment of attentional bias. The study incorporates two task paradigms—the DPT and the CTT—which may reflect different attentional processes, namely selective orienting toward negative stimuli and difficulty disengaging attention from such stimuli [18,19]. The protocol also compares two stimulus modalities—emotional faces and emotional words—which may differ in the extent to which they engage externally driven social-emotional processing or internally mediated semantic and self-referential processing [21,22]. In addition, the inclusion of cognitive function as a potential effect modifier is clinically important because cognitive control may influence emotional attention [16,17], and previous stroke research has suggested that cognitive status may affect the expression of attentional bias [13].

Several limitations should be acknowledged. First, because the present study uses a cross-sectional design, causal relationships between depressive symptoms and attentional bias cannot be established. Second, although this is a multicenter study, the participating hospitals are limited to a specific clinical context, and the eligibility criteria exclude patients with severe cognitive impairment, severe aphasia, impaired consciousness, and subarachnoid hemorrhage; therefore, the findings may not be generalizable to all patients with stroke. Third, task performance in the acute phase after stroke may be influenced by fatigue, fluctuations in physical condition, and environmental constraints during bedside assessment. Fourth, lesion location will be obtained from medical records rather than from detailed neuroimaging analyses, which may limit interpretation of the neurobiological correlates of attentional bias. Finally, depressive symptoms will be assessed using the HADS-D, which is a screening instrument rather than a diagnostic tool for major depressive disorder [25–27]. Accordingly, participants classified into the depressive symptom group in this study should be interpreted as individuals with elevated depressive symptoms rather than as patients with a formal psychiatric diagnosis.

Despite these limitations, the present study is expected to provide clinically relevant information for the design of early psychological interventions targeting depression-related cognitive processes after stroke. The findings may help identify which patients are most appropriate for future ABM-based interventions and which task and stimulus configurations should be prioritized in subsequent confirmatory studies. Effect size estimates derived from this study may also inform sample size planning for future longitudinal studies and randomized controlled trials. Existing evidence from randomized and meta-analytic studies in depressive populations supports the rationale for this line of investigation [11,12].

## Dissemination

The findings from this study will be disseminated through publication in a peer-reviewed international journal and presentation at relevant national and international academic conferences. A summary of the study findings may also be shared with participating institutions in accordance with institutional policies and ethical requirements. Any substantial protocol amendments that affect the scientific content of the study will be documented and updated in the study registration record, as appropriate.

## Data availability

Deidentified data underlying future study findings will be made available in an appropriate repository or upon reasonable request, subject to institutional ethics requirements and data protection regulations. Variables that could compromise participant privacy will not be shared publicly without appropriate safeguards.

## Acknowledgments

The authors thank the physicians, occupational therapists, and other medical staff at the participating institutions for their generous support in the preparation and conduct of this study. The authors also thank the patients who agree to participate in this research and contribute their valuable time and effort.

## Notes

### Competing Interest Statement

The authors have declared no competing interest.

### Clinical Trial

UMIN000059166

### Funding Statement

Yes

